# Comorbidity Risk Factors Contributing to COVID-19 Related Deaths in Florida, March 1, 2020-January 16, 2021

**DOI:** 10.1101/2021.04.14.21255434

**Authors:** Ursula K. Weiss, Jason D. Maynard, Katherine McDaniel, Alyssa Cohen, Marie Bailey, Scott A. Rivkees

## Abstract

**Objectives:** To assess the association of specific comorbid conditions to COVID-19 deaths in Florida among decedents 16 to 64 years of age.

**Methods:** This report uses Florida vital statistics death data over the period of March 1, 2020 through January 16, 2021, to estimate the effects of comorbid conditions on COVID-19 mortality for decedents 16 to 64 years of age. All cases of COVID-19 death occurring in Florida, regardless of resident status, were evaluated. The comorbidities, or contributing causes of death, identified in this report include Down syndrome, asthma, diabetes, pulmonary fibrosis, obesity, dementia, immunodeficiency, kidney disease, chronic obstructive pulmonary disease, hypertension, heart disease, and chronic liver disease and cirrhosis. The study uses a binary logistic regression to examine the relationship between COVID-19 and non-COVID-19 death and contributing causes of death based on information in the death record. Odds ratios were calculated as a residual of the logistic regression.

**Results:** Among COVID-19 deaths, Down syndrome was 15.26 times more likely to be a contributing cause of death compared to non-COVID-19 deaths followed by asthma (OR 7.74), diabetes (OR 6.11), pulmonary fibrosis (OR 5.13), obesity (OR 4.66), dementia (OR 4.51), immunodeficiency (OR 2.49), and kidney disease (OR 2.13). Chronic liver disease and cirrhosis (OR 0.95) and cancer (OR 0.79) had lower odds of being a contributing cause of death.

**Conclusions:** Heart disease, chronic liver disease and cirrhosis, and cancer were not risk factors for death from COVID-19 among decedents. Additional studies are needed to elucidate associations between race/ethnicity, socioeconomic status, and behavioral factors.

## Introduction

Underlying medical conditions and older age predispose individuals to adverse outcomes from coronavirus disease 2019 (COVID-19) infections *(1, 2)*. Several medical conditions associated with increased death from COVID-19 have been identified *(1, 2, 3)*; however, their association to COVID-19 as an underlying cause of death remains to be further defined. Such information can guide medical care and COVID-19 prevention strategies.

Florida is the third most populous state with more than 21.6 million residents *(4)*. Per-capita, Florida ranks second (20.5%) after Maine in those 65 years of age and older *(4,5)*. In addition, there are several million Floridians with underlying medical conditions *(6)*, which might make them at risk for severe health outcomes from COVID-19.

As of January 16, 2021, there have been 1,571,279 individuals who tested positive for COVID-19 in Florida *(7)*. 67,997 Florida residents have been hospitalized and 24,515 individuals have died with COVID-19 listed as either the underlying (primary) cause or a contributing factor *(7)*. Of those individuals who died from COVID-19 as the underlying cause of death, 83% were 65 years of age or older. In comparison with other states, per-capita, Florida ranks 29th in terms of number of cases and 26th in number of deaths *(8)*.

## Methods

This report uses Florida vital statistics death data over the period of March 1, 2020 through January 16, 2021, to estimate the effects of comorbid conditions on COVID-19 mortality. For the purpose of this study, contributing causes of death were examined to determine existing comorbidities. All cases of COVID-19 death occurring in Florida, regardless of resident status, where COVID-19 was listed as the underlying cause of death on the death certificate were evaluated. The population studied was limited to decedents between 16 to 64 years of age to mitigate the effects of older age and associated health outcomes. Furthermore, to assess potential high-risk age groups with a contributing cause of death, the sample was broken down by 5-year age increments. Records where COVID-19 was identified as the underlying cause of death by medical examiners (n=3,745) were compared with records where COVID-19 was not the underlying cause (n=47,670).

The study used a binary logistic regression to examine the relationship between COVID-19 and non-COVID-19 as the underlying cause of death and defined comorbidities or contributing causes of death based on information available in the death record. Comorbidities included in this analysis may overlap, as many adults may have more than one chronic condition. Comorbidities were selected based on existing COVID-19 published research, which suggests these conditions occurred prior to the COVID-19 underlying cause of death. Pneumonia and acute respiratory distress syndrome were excluded from the odds ratio analysis due to being possible symptoms of COVID-19. The contributing causes of death identified based on ICD-10 cause of death coding^*^ in this report included Down syndrome, asthma, diabetes, pulmonary fibrosis, obesity (BMI greater than or equal to 30), dementia, immunodeficiency, kidney disease, chronic obstructive pulmonary disease, hypertension, heart disease, and chronic liver disease and cirrhosis.

Odds ratios were calculated as a residual of the logistic regression. A simple random sample of the non-COVID-19 group was used to create a 10:1 ratio sample size, non-COVID-19 to COVID-19, respectively. The 10:1 ratio provided a stable model with the optimal c-statistic. SAS (version 9.4; SAS Institute) was used to conduct all analyses.

## Results

Table 1 displays deaths where COVID-19 was the underlying cause. Down syndrome was 15.26 times more likely to be a contributing cause of death as compared to non-COVID-19 deaths. In addition, decedents with asthma (OR 7.74), diabetes (OR 6.11), pulmonary fibrosis (OR 5.13), obesity (OR 4.66), dementia (OR 4.51), immunodeficiency (OR 2.49), and kidney disease (OR 2.13) had greater odds to be a contributing cause in deaths when COVID-19 was an underlying cause than non-COVID-19 deaths. Chronic liver disease and cirrhosis (OR 0.95) and cancer (OR 0.79) had lower odds of being a contributing cause of death. Table 2 shows for each contributing cause of death, the age distribution of individuals where COVID-19 was the underlying cause. For instance, among the 28 individuals with Down syndrome as a contributing cause of death, 32.1% (9 individuals) were ages 55-59 years. Overall, for each contributing cause of death examined, more deaths occurred in ages 55-64 years compared to all other age groups.

**Table 1.** Contributing Cause of Death Odds Ratios and 95% Percent Confidence Intervals in the 16-64 Age Group with COVID-19 as the Underlying Cause of Death Among Recorded Deaths in Florida, March 1, 2020 – January 16, 2021*

| **Contributing Cause of Death** | **Odds Ratio** | **(95% CI)** |
| --- | --- | --- |
| Down Syndrome | 15.262 | (8.382 - 27.787) |
| Asthma | 7.735 | (5.818 - 10.283) |
| Diabetes | 6.108 | (5.551 - 6.72) |
| Pulmonary Fibrosis | 5.127 | (3.302 - 7.961) |
| Obesity | 4.661 | (4.158 - 5.225) |
| Dementia | 4.507 | (3.013 - 6.741) |
| Immunodeficiency | 2.489 | (1.828 - 3.389) |
| Kidney Disease | 2.133 | (1.823 - 2.496) |
| Chronic Obstructive Pulmonary Disease | 1.26 | (1.084 - 1.465) |
| Hypertension | 1.183 | (1.057 - 1.323) |
| Heart Disease | 1.01 | (0.932 - 1.094) |
| Chronic Liver Disease and Cirrhosis | 0.945 | (0.741 - 1.205) |
| Cancer | 0.788 | (0.654 - 0.95) |

\* Wald chi-square test, p-value < 0.05 for Down syndrome, asthma, diabetes, pulmonary fibrosis, obesity, dementia, immunodeficiency, kidney disease, chronic obstructive pulmonary disease, hypertension, and cancer. *Note:* Individuals may be included in more than one contributing disease or condition category.

**Table 2.**
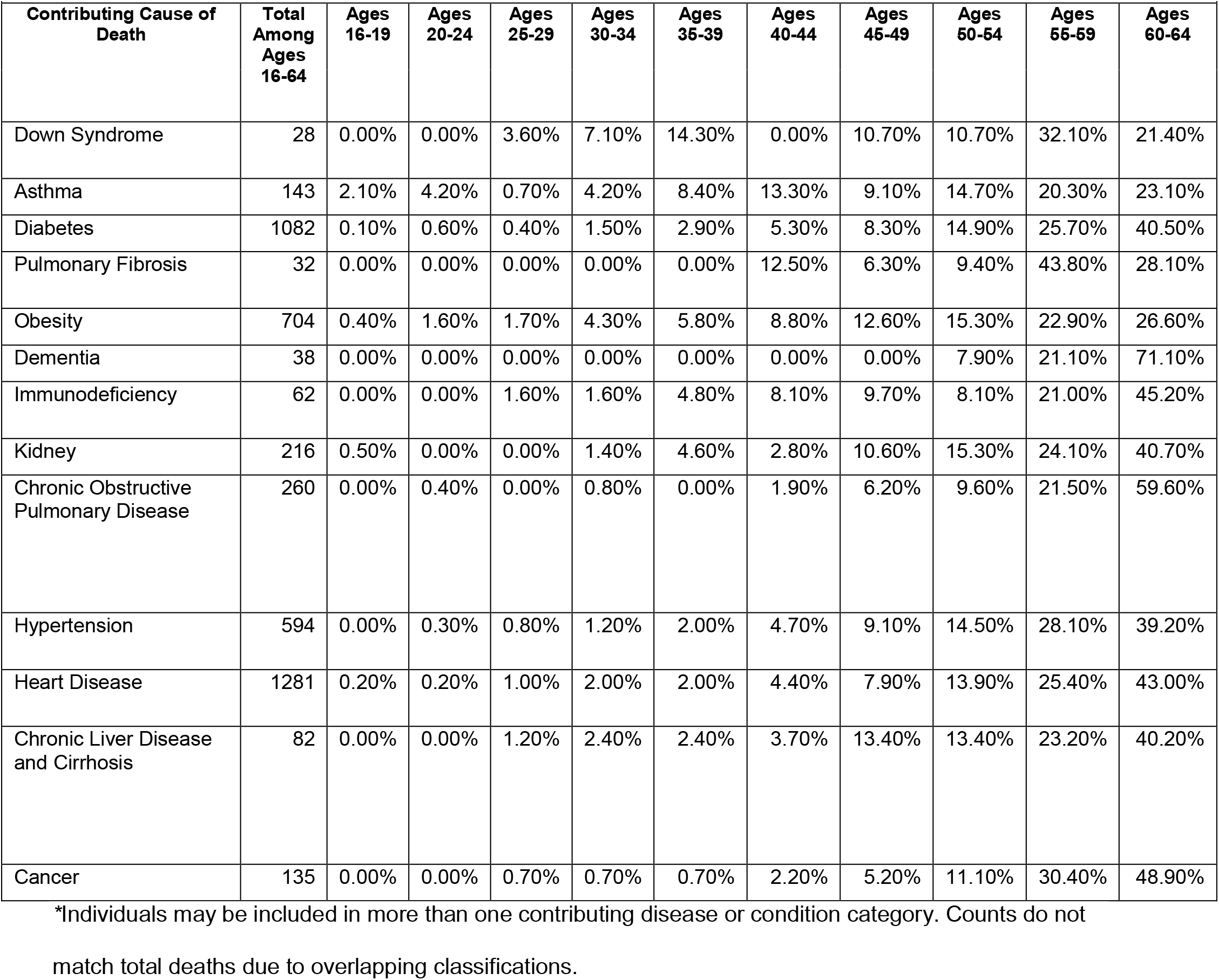
Distribution of Contributing Cause of Death by Age Group when COVID-19 is the Underlying Cause of Death Among Recorded Deaths in Florida, March 1, 2020 – January 16, 2021*

|  |  |  |  |  |  |  |  |  |  |  |  |
| --- | --- | --- | --- | --- | --- | --- | --- | --- | --- | --- | --- |
| **Contributing Cause of Death** | **Total Among Ages 16-64** | **Ages 16-19** | **Ages 20-24** | **Ages 25-29** | **Ages 30-34** | **Ages 35-39** | **Ages 40-44** | **Ages 45-49** | **Ages 50-54** | **Ages 55-59** | **Ages 60-64** |
| Down Syndrome | 28 | 0.00% | 0.00% | 3.60% | 7.10% | 14.30% | 0.00% | 10.70% | 10.70% | 32.10% | 21.40% |
| Asthma | 143 | 2.10% | 4.20% | 0.70% | 4.20% | 8.40% | 13.30% | 9.10% | 14.70% | 20.30% | 23.10% |
| Diabetes | 1082 | 0.10% | 0.60% | 0.40% | 1.50% | 2.90% | 5.30% | 8.30% | 14.90% | 25.70% | 40.50% |
| Pulmonary Fibrosis | 32 | 0.00% | 0.00% | 0.00% | 0.00% | 0.00% | 12.50% | 6.30% | 9.40% | 43.80% | 28.10% |
| Obesity | 704 | 0.40% | 1.60% | 1.70% | 4.30% | 5.80% | 8.80% | 12.60% | 15.30% | 22.90% | 26.60% |
| Dementia | 38 | 0.00% | 0.00% | 0.00% | 0.00% | 0.00% | 0.00% | 0.00% | 7.90% | 21.10% | 71.10% |
| Immunodeficiency | 62 | 0.00% | 0.00% | 1.60% | 1.60% | 4.80% | 8.10% | 9.70% | 8.10% | 21.00% | 45.20% |
| Kidney | 216 | 0.50% | 0.00% | 0.00% | 1.40% | 4.60% | 2.80% | 10.60% | 15.30% | 24.10% | 40.70% |
| Chronic Obstructive Pulmonary Disease | 260 | 0.00% | 0.40% | 0.00% | 0.80% | 0.00% | 1.90% | 6.20% | 9.60% | 21.50% | 59.60% |
| Hypertension | 594 | 0.00% | 0.30% | 0.80% | 1.20% | 2.00% | 4.70% | 9.10% | 14.50% | 28.10% | 39.20% |
| Heart Disease | 1281 | 0.20% | 0.20% | 1.00% | 2.00% | 2.00% | 4.40% | 7.90% | 13.90% | 25.40% | 43.00% |
| Chronic Liver Disease and Cirrhosis | 82 | 0.00% | 0.00% | 1.20% | 2.40% | 2.40% | 3.70% | 13.40% | 13.40% | 23.20% | 40.20% |
| Cancer | 135 | 0.00% | 0.00% | 0.70% | 0.70% | 0.70% | 2.20% | 5.20% | 11.10% | 30.40% | 48.90% |

\*Individuals may be included in more than one contributing disease or condition category. Counts do not match total deaths due to overlapping classifications.

## Discussion

These data confirm that several underlying medical conditions are associated with COVID-19 deaths in individuals less than 65 years of age. We also define the odd ratios of a number of these conditions contributing to death. Of the conditions evaluated, individuals with Down syndrome, asthma, diabetes, and pulmonary fibrosis had the highest likelihood of death, followed by those with obesity, dementia, immunodeficiency, and kidney disease. Differing with the observation of others *(1, 2, 9)*, we did not find that heart disease, chronic liver disease and cirrhosis, nor cancer were risk factors for death from COVID-19 among decedents. In fact, the study shows that for all non-COVID-19 deaths, cancer was more likely to be a contributing cause. Moreover, in terms of age as an additional factor, the data shows that most deaths occur in individuals ages 55-64 years.

The findings in this report are subject to some limitations. In Florida, coding of underlying and contributing causes of death are conducted by the National Center for Health Statistics. Thus, there could be a delay in the reporting of COVID-19-related deaths. Odds ratio analysis was conducted on decedents between 16-64 years or age, which might skew results to those contributing causes of death more closely associated with older decedents. Also, the 10:1 ratio used for this study produced slight over-dispersion, meaning variance was slightly higher than it would have been with a smaller ratio. Our analysis does not consider the impact of racial and ethnic disparities on contributing causes associated with COVID-19-related deaths and does not consider geographic differences such as proximity to hospitals, which could also impact contributing causes associated with COVID-19-related deaths.

## Conclusion

Despite limitations, this report provides important data for comorbid conditions as a contributing factor for deaths where COVID-19 is the underlying cause. Such information is important in guiding medical care and public health COVID-19 prevention policies. Additional studies are needed to elucidate associations between race/ethnicity, socioeconomic status, and behavioral factors. Regional and state-level efforts to more thoroughly examine the association of comorbidities with COVID-19-related deaths could lead to targeted, community-level and individual-level mortality prevention initiatives *(3)*.

## Data Availability

The datasets generated during and/or analyzed during the current study are not publicly available due to HIPPA and PHI privacy but may be available from the Florida Department of Health, Division of Public Health Statistics and Performance Management through appropriate data use agreements on reasonable request.

## Footnotes

* ICD-10 cause of death codes for the causes analyzed in the study: Down Syndrome – Q90.0, Q90.1, Q90.2, Q90.0; Asthma – J45-J46; Diabetes – E10-E14; Pulmonary Fibrosis-J84; Obesity – E66; Dementia – F03; Immunodeficiency – B20-B24, D80-D84; Kidney Disease – N00-N07, N17-N19, N25-N27; Chronic Obstructive Pulmonary Disease – J40-J44; Hypertension – I10; Heart Disease – I00-I09, I11, I13, I20-I51; Chronic Liver Disease and Cirrhosis – K73-K74, K70; Cancer – C00-C97.

